# Validation of rectal swabbing for total and aerobic gut microbiota study

**DOI:** 10.1101/2024.07.22.24310623

**Authors:** Julie Marin, Paul Albin Bertoye, Andre Birgy, Samira Dziri, Mathilde Lescat

## Abstract

In microbiota research, whole stool sampling is the conventional approach but can be problematic or infeasible for certain patients. This study aims to validate the use of rectal swabbing as an alternative method for microbiota analysis and determine optimal storage conditions suitable for various clinical settings, including intensive care units. We evaluated different sampling techniques, conservation media and storage temperatures. Our findings indicated that rectal swabs yield microbiota diversity comparable to whole stool samples. Notably, storage conditions significantly impacted microbiota profiles, with increased *E. coli* and *Enterococcus sp*. quantifications observed at room temperature. Consequently, we recommend immediate refrigeration of rectal swabs to reliably assess aerobic and total microbiota, particularly for patients requiring urgent care such as antibiotic treatment.

**Importance:** We developed a pragmatic approach to study total and aerobic gut microbiota, applicable in numerous clinical units such as intensive care or emergency units, where whole stool sampling is often impractical. This approach employs ESwab™ devices which are already commonly used in hospital.

## Introduction

The human microbiota, a diverse community of microorganisms residing in our bodies, is a key area in health research. Particularly, the gut microbiota, mainly influenced by diet and antibiotics, plays a vital role in human health (De Filippo et al. 2010; Dethlefsen and Relman 2011). Dysbiosis, characterized by imbalances in microbial composition, has been linked to various diseases, including obesity, inflammatory bowel diseases and cirrhosis (Liu et al. 2012; Tremaroli and Bäckhed 2012; Xu et al. 2012; Purchiaroni et al. 2013). Quantitative assessment of aerobic bacteria is also crucial for understanding severe infections. Indeed, imbalances in the gut microbiota are linked to infections, such as urinary tract infections caused by *Escherichia coli*, which correlate with elevated *E. coli* quantities in the digestive tract pre-infection. Understanding the interactions between the microbiota and pathogenic agents is of primary importance in the prevention and treatment of multiple diseases, paving the way for novel therapeutic strategies targeting microbiota modulation.

Investigating the gut microbiota in numerous patients presents challenges, especially when it comes to obtaining high-quality samples through conventional total fecal sampling methods. This difficulty is particularly pronounced in severely ill patients who require urgent antibiotic therapy. A comparison between rectal swabs and whole stool sampling has shown promise as an alternative, yielding comparable microbial compositions using 16S rRNA analysis (Radhakrishnan et al. 2023). However, these studies utilized dry swabs, which are no longer standard in microbiology analyses. Previous evaluations of gut microbiota using FecalSwab™ highlighted some differences compared to analyses performed on whole stools stored without a conservation medium. Notably, no evaluations have been conducted using ESwab™ devices which are the most often swabs used in hospitals (Tedjo et al. 2015a).

Concerning, culturomic based analysis other researchers have also investigated the use of rectal swabs, particularly ESwab™, which maintains the viability of several aero-anaerobic bacteria such as *Streptococcus* pneumoniae and *Haemophilus influenzae*, as well as strictly anaerobic bacteria, for up to 48 hours at room temperature, with even better preservation at refrigerated temperatures (Van Horn et al. 2008). Additionally, comparative studies between ESwab™ and FecalSwab™ media have been conducted to assess the viability of various diarrheagenic bacteria under different temperature conditions. These studies found that FecalSwab™ devices provided better viability for some diarrheagenic strains and reference strains, such as *E. coli* ATCC 25922 and *Enterococcus faecalis* ATCC 29212, under refrigerated and frozen conditions (Hirvonen and Kaukoranta 2014). However, these studies did not focus on the viability of various natural isolates of commensal *Enterobacteriaceae* or *Enterococcus species*, which are the most frequent bacteria involved in infections originating from the digestive tract in cirrhotic patients. Beyond bacterial culture with reference strains, other studies have demonstrated that the ESwab™ medium can reliably be used to study the vaginal or skin microbiota (Bjerre et al. 2019; Mattei et al. 2019). Thus, the validation of rectal swabs using ESwab™, (the devices we use in the hospital), as a reliable tool for microbiota studies (16SrRNA and cutluromic analysis) is crucial.

To address the challenges of studying microbiota diversity in intensive care and other emergency units, we aimed in this study to (i) validate the sampling conditions and (ii) establish a robust conservation methodology using rectal ESwab™ for analyzing gut microbiota, encompassing both total and aerobic populations.

## Materials and methods

### Healthy volunteer sampling

Thirteen healthy donors provided fecal samples for our study. None had taken antibiotics or experienced any medical issues in the preceding 6 months. In accordance with APHP ethics regulations and the Declaration of Helsinki, all participants received comprehensive information and provided consent. Fecal samples and rectal swabs, self-collected by the volunteers, were directly placed into sterile containers or ESwab™. Subsequently, the samples were transported to the laboratory within 24 hours of collection (**Figure 1A**). The figure 1 (A and B) was made with BioRender (biorender.com).

**Figure 1.**
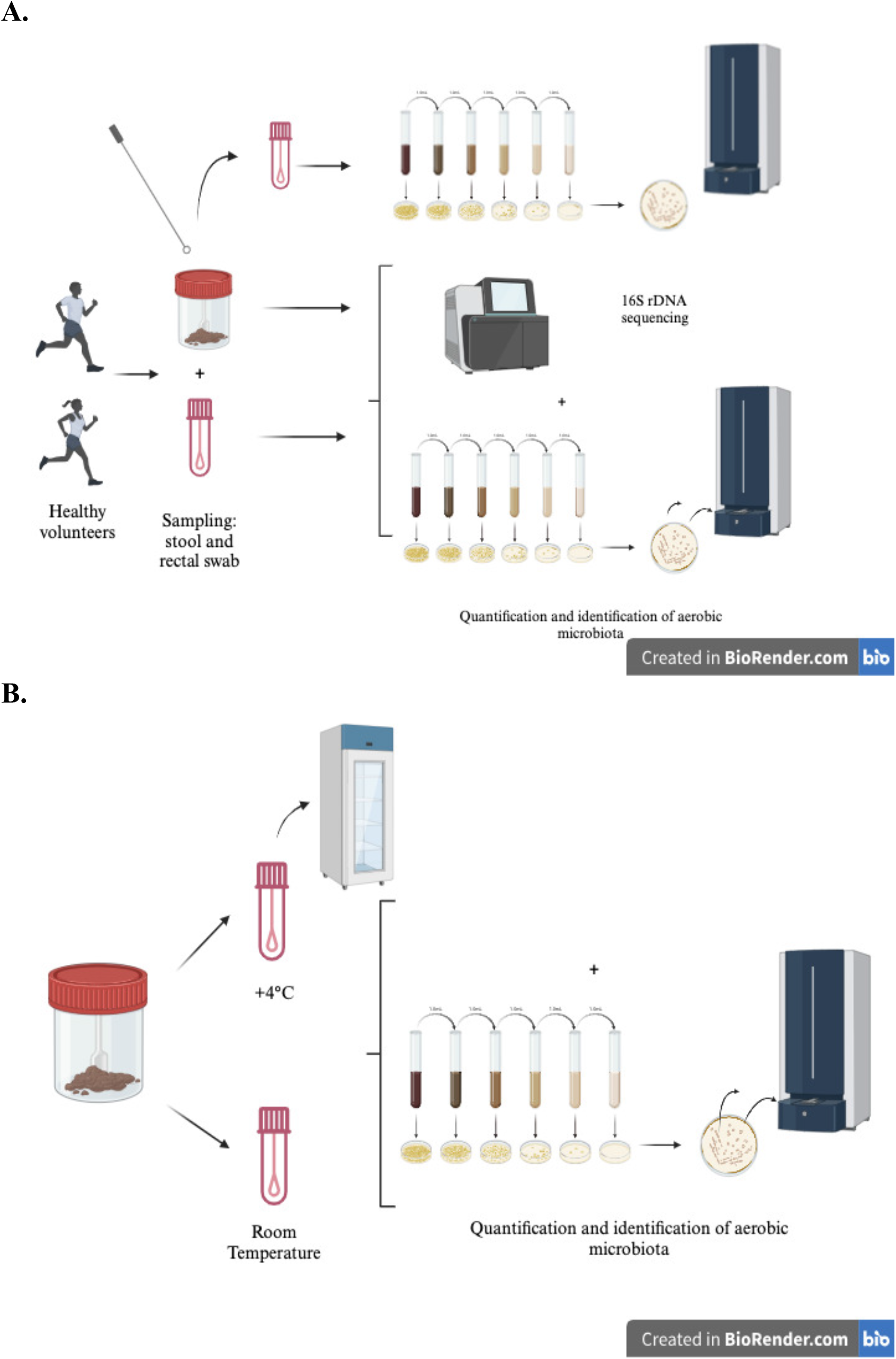
Schematic description of the protocols tested to validate (A) the conditions of sampling (total stool/rectal swab) in ESwab™ medium and (B) the temperature condition of storage for the total and aerobic microbiota studies in intensive conditions of clinical practice.

### Evaluation of the rectal swab method for total microbiota sequencing and analysis

#### DNA Extraction

We collected 200 mg of total feces or the pellet of liquid contained in the ESwab™ (Copan Diagnostics, Italy) devices after centrifugation during 10 mn at 5000 rpm. We then used the DNeasyPowerSoil Pro 250kit® (Qiagen, Les Ulis, France) for total manual DNA extraction, following the manufacturer’s protocol.

#### 16SrRNA Gene Sequencing

Intestinal gut microbiota composition in collected samples was analyzed using 16SrRNA gene sequencing (Yarza et al. 2014) targeting the V4 variable region (Bukin et al. 2019). The manufacturer’s instructions for the library preparation were followed, using KAPAHiFi HotStart ReadyMix (RocheLaboratories, Basel, Switzerland). Briefly, the amplification of the 16S region (“PCR1”) was performed with the 515 forward primer (5′-GTGCCAGCMGCCGCGGTAA-3′) and the 806 reverse primer (5′-GGACTACHVGGGTWTCTAAT-3′) as described elsewhere (Caporaso et al. 2011). The multiplexing step of *the* samples on the16S amplified regions (“PCR2”) required the Nextera® Index Kit and Nextera®XT Index Kit V2 SetD. PCRs products were quantified using the Qubit HS® normalized to 4nM for pooling and sequencing.16SrRNA gene sequencing was performed with the Illumina MiSeq technology (paired-end reads 2*250bp).

#### Pipeline settings

We followed the pipeline described in Li et al. (Li et al. 2016) the main steps are below. We controlled the quality and removed ambiguous sequences with FastQC (Andrews, 2010). We trimmed the residual sequences with Timmomatic (Bolger et al. 2014) (base quality score >20 and read lengths >50bp). We sorted the filtered reads into fragments 16 rRNA with SortMeRNA (Kopylova et al. 2012) using the rRNA reference database SILVA_SSU (Quast et al. 2013). Next with mothur (Schloss 2020) we i) removed sequences with ambiguous bases and sequences without overlapped regions (longer than 310 bp), ii) aligned the bacterial 16S rRNA sequences to the regionally enriched bacterial references, iii) clustered highly similar sequences (with one or two nucleotide differences), iv) identified and removed chimeric sequences (UCHIME (Edgar et al. 2011)), v) taxonomically assigned the sequences to different phylotypes using a naive Bayesian algorithm (Wang et al. 2007) and v) determined the taxonomic rank and computed the relative abundance for each phylotype.

#### Diversity analysis

To be able to compare alpha-diversity estimators among samples and methods, all samples were standardized to the same number of sequences (1000 sequences) by randomly resampling the OTU table (‘rarefy_even_depth’ function in the R package phyloseq(McMurdie and Holmes 2012)). We conducted alpha-diversity analysis at the bacterial genus level. We computed the Shannon index, which takes into account the number of OTU and their abundances (‘estimate_richness’ function in the R package phyloseq(McMurdie and Holmes 2012)). Finally, we evaluated the OTU differential abundance (‘DESeq2’ function in the R package DESeq (Love et al. 2014).

### Evaluation of the rectal swab method for aerobic microbiota study

#### Comparison of sampling strategies and evaluation of ESwab™ medium

To compare sampling conditions among 13 healthy volunteers, we estimated bacterial quantification by plating dilutions of suspensions from freshly weighed feces in physiological serum (as the reference) or suspensions from freshly weighed feces in ESwab™ (Copan Diagnostics, Italy), as well as from weighted rectal swabs ESwab™ for each individual. These were plated on UriSelect4 plates (Bio-Rad, La Coquette Marches, France), as detailed in **Figure 1A**. Aerobic microbiota analysis involved quantifying bacteria in four main classes: *Escherichia coli, Enterococcus species* and other Gram negative and Gram positive bacteria species. Main morphologies were confirmed using Matrix assisted Laser Desorption Ionisation Time of Flight (MALDI-TOF) mass spectrometer (Bruker, Champs-sur -Marne, France) identification before classification into the aforementioned groups. For each condition (WSeS for whole stool in ESwab™, RSeS for rectal swab in ESwab™

#### Evaluation of Temperature storage

To validate the storage temperature conditions post-sampling, we conducted a comparison of *E. coli* and *Enterococcus sp* quantification after inoculating feces from 10 other healthy volunteers into two tubes of ESwab™. One tube was stored at +4°C while the other was kept at room temperature. For each tube, we quantified *E. coli* and *Enterococcus sp*. at various time points: initial time (T0), 2 hours (T+2), 4 hours (T+4), 8 hours (T+8), and 24 hours (T+24). Quantification involved plating dilutions of the ESwab™ medium from each feces onto UriSelect4 plates (Bio-Rad, Marne La Coquette, France), as outlined in **Figure 1B**. In each case, suspected colonies of *E. coli* and *Enterococcus sp* were confirmed using MALDI-TOF identification (Bruker, Champs-sur -Marne, France). Then, for each condition of temperature (+4°C and 21°C) at each time point (T0, T+2; T+4, T+8 and T+24), we calculated the ratio of logarithms of quantifications of *E. coli* and *Enterococcus sp*, divided by the logarithm of the corresponding quantification at T0, respectively.

## Results

### Validation of the sampling procedure: Evaluation of the rectal swab method for total microbiota study

We compared the alpha and beta diversities obtained with two sampling methods, rectal swab and whole stool, and two conservation media, ESwab™. We did not find any difference for the alpha diversity (OTU richness or Shannon index) between the two sampling methods, whether with ESwab™ conservation (paired Wilcoxon test, pvalue=0.25 and 0.84 for OTU richness and Shannon index respectively) (**Figure 2**). Next, we tested the OTU differential abundance. With the ESwab™ conservation method, we did not find any difference, with the exception of the genus *Enterococcus* which was overrepresented in the whole stool samples compared to the rectal swab samples (Wald test, pvalue=9.88e-06).

**Figure 2.**
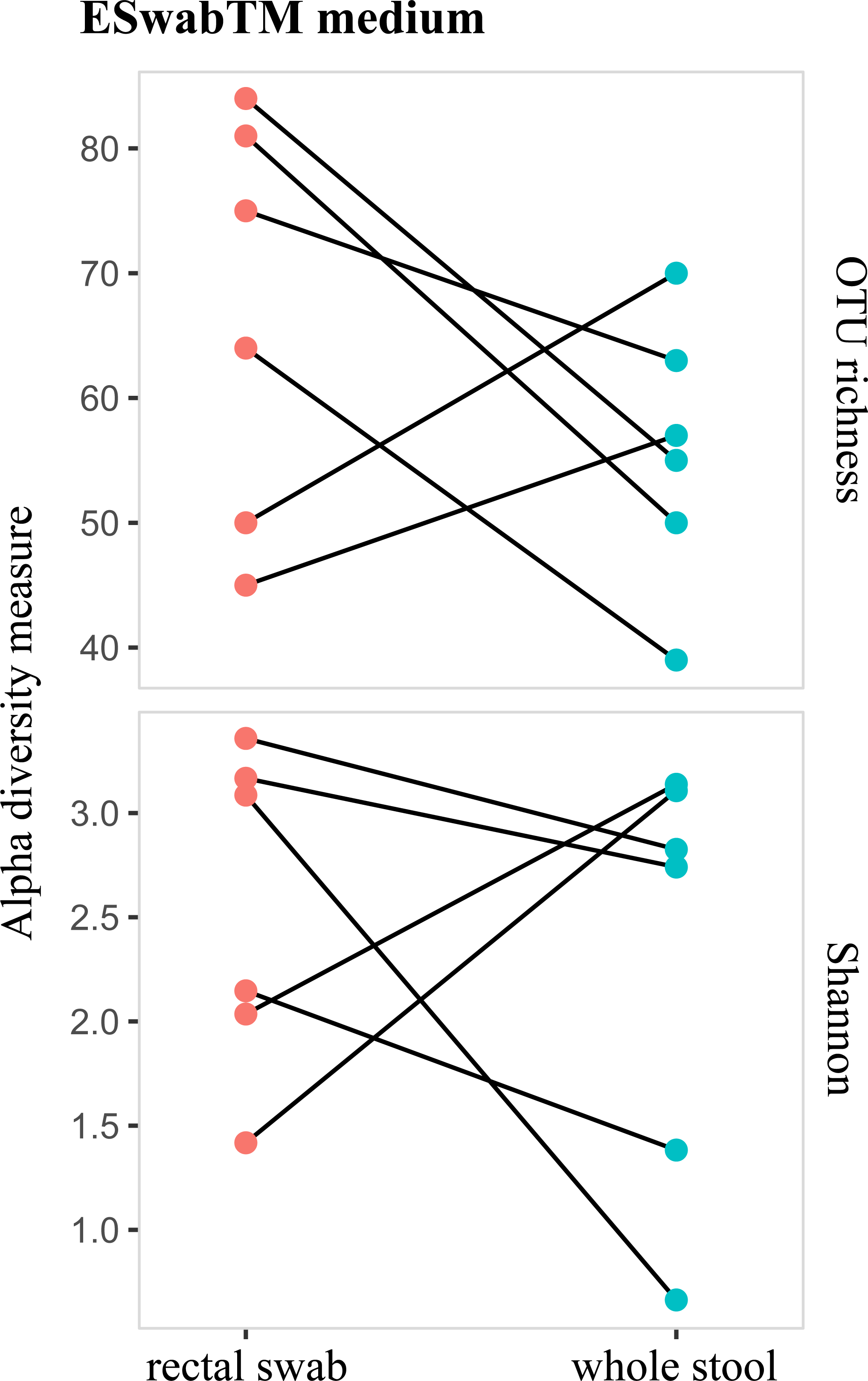
Comparison of the alpha diversity (OTU richness and Shannon index) among sampling (rectal swab versus whole stool) in ESwab™.

### Evaluation of the rectal swab method for an aerobic microbiota study

Overall, for the 13 volunteers, 11 brought back complete sampling (whole stool and rectal swabs), 2 only brought back whole stool. We then inoculated whole stool in ESwab™ medium and compared the quantifications of the 4 main class of species between the whole stool, we considered as a reference, the whole stool in ESwab™ to determine the effect of the medium and the rectal swab in ESwab™ to determine the effect of the rectal swabbing. Overall, we observed a conservation of all Gram negative bacteria (*E. coli* and other when present in whole stool) in all conditions. However, we detected an important increase of *E. coli* quantifications in RSeS (multiple t-test with Benjamini-Hochberg correction, pvalue = 0.036 and 0.042 when comparing RSeS to WS and RSeS respectively). For Gram positive bacteria, we found at least the species *Enterococcus* in their whole stool. We observed a loss of one and two out of the 13 donors in ESwab™ medium (for WTeS and RSeS respectively). We also observed the effect of rectal swabbing with the loss of respectively one *Enterococcus sp* and other Gram positive bacteria (*Streptococcus sp)* for respectively one and three volunteers (**Figure 3, Table S1**).

**Figure 3.**
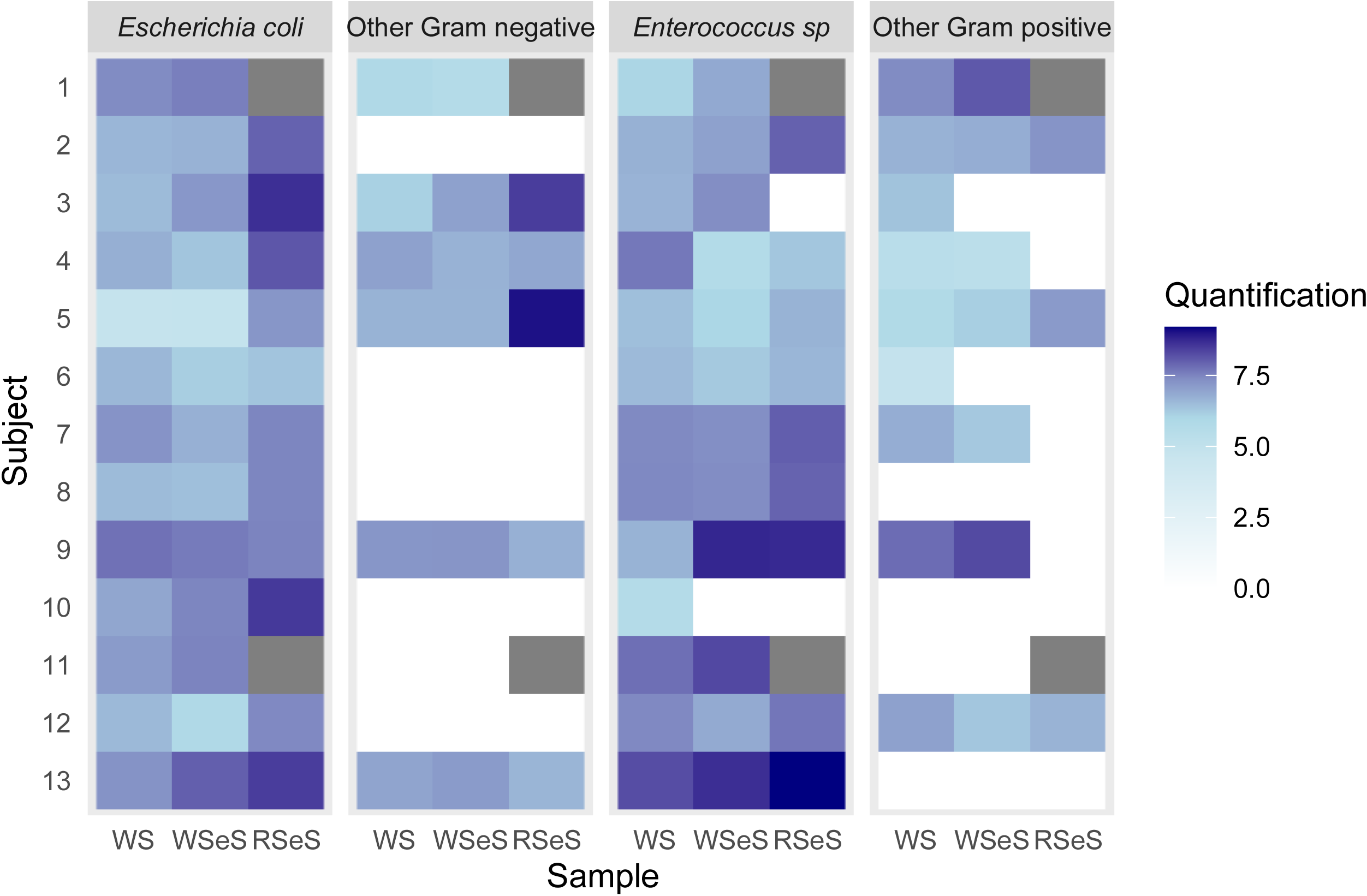
Quantifications of four main groups of bacteria (*Escherichia coli*, other Gram negative bacteria, *Enterococcus sp*. and *other* Gram positive bacteria) for different media and sampling procedure. The quantifications were in logarithmic scales. The different conditions were indicated respectively as WSeS for whole stool in ESwab™ and RSeS for rectal swab in ESwab™. Missing samples are indicated in grey.

Considering increase of *E. coli* quantification in rectal swabs and the fact that healthy volunteers brought their samples (whole stool without any medium and rectal swabs in ESwab™ medium) without any precaution of storage, we hypothesized that that media could have enhance multiplication of some bacteria in it. We then performed a comparison of *E. coli* and *Enterococcus sp* quantifications after inoculating feces from 10 other healthy volunteers into two tubes of ESwab™ in two conditions of temperature of storage (+4°C and room temperature) and quantified those two types of bacteria through time (T0, T+2, T+4, T+8, and T+24h) (**Figure 4, Table S2**). We observed a significant increase of quantifications of *E. coli* at room temperature (Pearson’s correlation test, r = 0.62, pvalue = 1.65e-06) after 4 hours of incubation. We also detected an increase, although non-significant of *Enterococcus sp*. quantifications at room temperature (Pearson’s correlation test, r = 0.25, pvalue = 9.24e-02). Quantifications remained stable at +4°C for both quantifications of *E. coli* and *Enterococcus sp*. (Pearson’s correlation test, r = 0.04 and 0.07, pvalue = 0.78 and 0.63 respectively).

**Figure 4.**
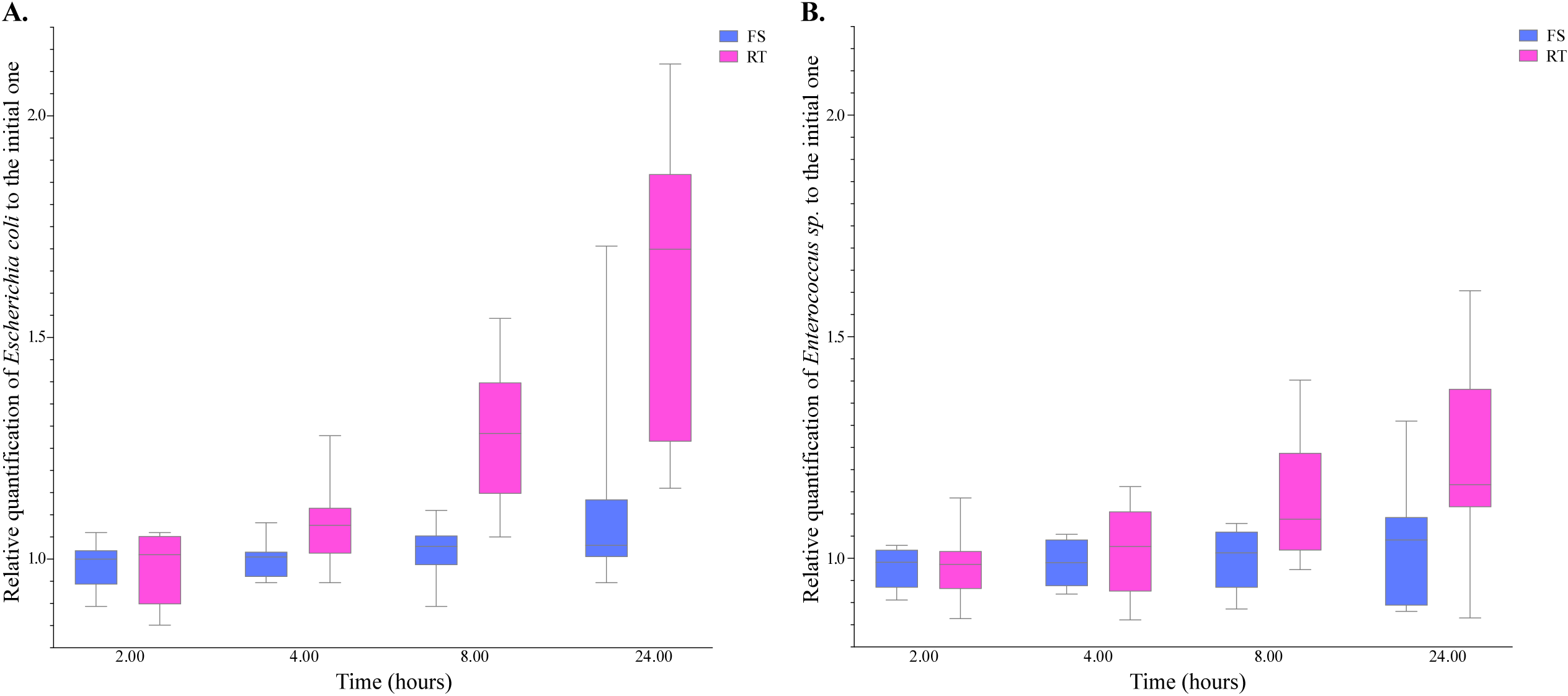
Relative quantifications of *Escherichia coli* and *Enterococcus sp* across time to the corresponding ones at T0. The quantifications were in logarithmic scales. FS and RT corresponded to the temperature conditions of storage, +4°C and room temperatures.

## Discussion

In this study, we aimed to validate the conditions of sampling and conservation methodology for microbiota (total and aerobic) using rectal ESwab™. Our goal was to develop a pragmatic approach applicable in numerous clinical units such as intensive care or emergency units, where obtaining whole stool samples from fragile patients is often impractical.

We validated the rectal swab and the use of ESwab™ medium method for microbiota analysis by comparing different sampling conditions and conservation media using samples from 13 healthy volunteers. This evaluation was never been done to our knowledge on natural isolates of aerobic bacteria from digestive origin (Van Horn et al. 2008; Tedjo et al. 2015b). The samples included both whole stool and rectal swabs. We focused on alpha-diversities obtained from total microbiota analysis and quantifications of aerobic bacteria by aerobic cultures across four main classes: *E. coli, Enterococcus sp*., other Gram negative and Gram positive bacteria. We observed a notable increase in *E. coli* quantification under rectal swabbing conditions. Healthy volunteers performed their swabbing and brought back it to the lab. They possibly did not properly conserve the sample at +4°C, which might explain our quantification results. To investigate this hypothesis, we tested storage conditions and found that samples stored at room temperature showed significant increases in *E. coli* quantifications, and in a lesser degree in *Enterococcus sp*. quantifications, while quantifications remained stable at 4°C.

This finding underscores the importance of maintaining appropriate storage temperatures for accurate bacterial quantifications.

Our findings also indicated a significant decrease in Gram positive retrieval for Gram positive bacteria such as *Enterococcus sp*. and *Streptococcus sp*. under rectal swabbing conditions and ESwab™. These observations suggest that rectal swabs might be less effective for certain bacterial populations, possibly due to challenges in conservation or sampling efficiency. Nevertheless, our study demonstrates that rectal swabs can reliably reflect the alpha diversity (OTU richness and Shannon index) observed in whole stool samples even if we observed some differences for *Enterococcus sp*. in quantifications and OTU richness. Differences in the relative abundance of bacteria species has also been reported when comparing total stool and Fecal transport swabs (FecalSwab™, Copan) (Tedjo et al. 2015). These differences might be explained by a slight growth disadvantage of certain bacterial taxa in some media, *Enterococcus species* in ESwab™ medium in our case.

## Conclusion

Overall, despite some variability in quantifications of specific bacterial populations, rectal swabs remain a viable method for microbiota studies, particularly when optimized storage conditions are maintained. This decision simplifies procedures in hospital settings where ESwab™ devices are already commonly utilized.

## Data Availability

All data produced in the present study are available upon reasonable request to the authors

## Ethical authorizations

All individuals were collected thanks to ethical authorization n°CLEA 2019-72 obtained by the “Comité local de Protection des Personnes” des hôpitaux universitaires de Paris Seine Saint Denis.

## Acknowledgements

We thank all the participants of the study.

## Supplementary Online Material (SOM)

**Table S1.** Quantifications of four main groups of bacteria (Escherichia coli, other Gram negative bacteria, *Enterococcus sp*. and other Gram positive bacteria) for different sampling procedure.

**Table S2.** Quantifications of *Escherichia coli* and *Enterococcus sp*. in ESwab™ medium across the time at fridge and room temperatures.

